# Association between lifestyle factors and weight in Japan university students during COVID-19 mild lockdown: a quantitative study

**DOI:** 10.1101/2023.05.24.23290491

**Authors:** Haruka Arimori, Norio Abiru, Shimpei Morimoto, Tomoya Nishino, Atsushi Kawakami, Akie Kamada, Masakazu Kobayashi

## Abstract

We investigated the lifestyle factors influencing weight gain in university students during restrictions (mild lockdown) imposed owing to the novel coronavirus disease pandemic in Japan. In this cross-sectional study, a questionnaire survey of Nagasaki University students undergoing health examinations was conducted in 2021. Students reporting ≥3 kg weight gain were included in the weight gain group; the remaining students were in the non-weight gain group. Fisher’s exact test and binary logistic regression were performed to detect the associations between weight gain and each lifestyle factor. We included 3,059 respondents (response rate: 45.7%), and 9.5% respondents reported ≥3 kg weight gain. The following factors were associated with weight gain (odds ratio, 95% confidence interval, p value from Fisher’s exact test): dining out for ≥4 times/week (2.16 [1.40, 3.32], p = 8.7 × 10^−4^), gaming time of ≥4 h/day (2.26 [1.45, 3.47], p = 2.4 × 10^−4^). Binary logistic regression among the four highest odds ratios showed that after adjusting for other factors frequently dining out and prolonged gaming time were significantly associated with weight gain. Prolonged gaming and frequently dining out were associated with weight gain in students during the mild lockdown.

## Introduction

The rapid increase in obesity is a global health issue. In the United States of America, the highest increase in the prevalence of obesity from 1991 to 1998 was noted among individuals aged 18–29 years (from 7.1% to 12.1%) [1]. According to a report by the Ministry of Health, Labour and Welfare of Japan, the prevalence of obesity was 23.1% among men in their 20 s, 29.4% among men in their 30 s, and 39.7% among men in their 40 s [2]. Kobayashi et al. reported that metabolic syndrome was observed in 3.3% of men and no women in their cohort study of university students [3]. These findings indicate the necessity of countermeasures against obesity in youth.

Following the novel coronavirus disease 2019 (COVID-19) outbreak caused by the severe acute respiratory syndrome coronavirus 2 in December 2019, the World Health Organization declared COVID-19 a pandemic on March 11, 2020. In several countries, curfews (lockdowns) were lawfully implemented and penalties applied. On April 7, 2020, the Japanese government declared a state of emergency, and to prevent the spread of infection, they requested individuals to refrain from nonessential and nonurgent outings, to avoid social contact to the maximum extent, and to stay at home to ensure social distancing. This non-enforceable and non-punitive request to stay indoors is hereafter referred to as “mild lockdown” in the present study. Although this restriction of activity was considered effective for controlling the spread of COVID-19 infection, it was accompanied with numerous adverse effects, such as lack of exercise, weight gain, behavioral addiction, lack of sun exposure, and social isolation; in particular, staying at home for several months led to changes in social habits, and there was a risk of changes in personal health [4,5].

In the youth, there were major lifestyle changes; particularly, in university students, learning at home and attending online lectures as well as the voluntary restriction of social club activities and restriction of part-time work caused several lifestyle changes in physical activity, diet, and sleep habits, which were associated with weight gain.

The mild lockdown will continue until 2022, and it is speculated that prolonged lifestyle changes will exacerbate weight gain and health issues among university students. Although young adulthood, such as the university years, is considered a crucial time for the development of metabolic abnormalities and increased obesity [6], the impact of lifestyle changes during the mild lockdown on student health has not adequately been examined.

In the present study, in order to identify the factors that contribute to weight gain in youth during mild lockdown, we investigated the relationship between weight gain and lifestyle changes during the mild lockdown in students at Nagasaki University and explored the lifestyle factors and their cutoffs associated with weight gain among youth in the face of a major lifestyle change of 1–2 years of mild lockdown.

## Materials and Methods

### Participants

The participants were students of Nagasaki University receiving student medical examinations in 2020 and 2021. Individuals who understood Japanese, irrespective of their nationality, and who consented to the present study were included as participants. The authors accessed personally identifiable information of the subjects during or after data collection.

In 2020, there were a total of 9,179 students at Nagasaki University; of them, 6,065 received a health examination, and 3,722 students completed a questionnaire (response rate, 61.2%). We excluded 1 student with an incomplete questionnaire; therefore, a total of 3,721 students (1,850 men) were included in the analyses.

In 2021, there were a total of 9,031 students at Nagasaki University; of these, 6,675 received a health examination, and 3,059 students (1,705 men) who were not first-year students, who consented to the study, and who completed the questionnaire response rate, 45.7%) were included in the analyses (S1 Fig).

### Study design

The present study was conducted with the approval of the ethical review board of Nagasaki University (approval number: 20062604) and in accordance with the principles of the Declaration of Helsinki.

This study is a cross-sectional single-center survey. The study period was from July 27 to November 27, 2020, and from April 20 to June 11, 2021. Researchers and research assistants visited Nagasaki University and requested voluntary participation from students to receive medical examinations in 2020 and 2021. Before implementing the study, written consent was obtained from eligible individuals. After obtaining consent in Japanese, participants were requested to complete an electronic questionnaire.

The online survey was distributed as an electronic questionnaire via Google Forms® (QR code provided), which is a social media service. Researchers and research assistants adopted infection prevention measures, such as handwashing and physical distancing, and used surgical masks, face shields, medical gloves, and sanitizing wipes. The questionnaire survey was conducted during waiting periods before and after the medical examinations. The time required to complete the questionnaire was approximately 5 minutes. The data was confidentially analyzed by the researchers.

To assess the lifestyle of university students, we newly developed our own questionnaire based on the Pittsburgh Sleep Quality Index [7] and the National Health and Nutrition Survey [2] of the Ministry of Health and Welfare. From the Pittsburgh Sleep Quality Index, we used bedtime, waking up time, and sleep duration (all listed), and from the National Health Survey, we used frequency of breakfast and frequency of dining out (all listed). The questionnaire included 17 items under 6 sections.

1. Personal information obtained via two questions (age and sex)
2. Subjective evaluation on the changes in their weight and the general aspects of their lifestyles obtained via two questions (weight change: “gain of ≥3 kg,” “gain of <3 kg,” “unchanged,” “loss of <3 kg,” and “loss of ≥3 kg”; and general lifestyle change: “changed greatly,” “changed a little,” and “unchanged”)
3. Information regarding physical activity obtained via three questions (time spent at home: h/day, frequency of part-time work: times/week, and frequency of social club activities: times/week)
4. Information regarding diet obtained via two questions (frequency of breakfast: times/week, frequency of dining out: times/week)
5. Information regarding daily rhythm obtained via three questions (bedtime, waking up time, and sleep duration: h/day)
6. Information regarding lifestyle obtained via five questions (smoking habit, drinking habit: times/month, alcohol amount: units/time, gaming time: h/day, and internet surfing time: h/day)

We examined students who responded that they observed a weight gain of ≥3 kg (WG group) and the remaining students for all other categories (non-WG group). For each category in each lifestyle factor in sections (3)–(6), categorical variables were stratified into categories of 3–4 groups for before and during the mild lockdown. Further, the change was dichotomized to evaluate whether the change within a participant between before and after the lockdown crossed a threshold between adjacent categories.

In section (2), questions about changes in weight and lifestyle were asked before and during mild lockdown in both 2020 and 2021, respectively: in 2020, before: March–June 2019, after: from March 2020 to during survey (July–November 2020); in 2021, before: October–December 2019, and later: October–December 2020. For each lifestyle factor in sections (3)–(6), we questioned the daily lifestyle before and during the mild lockdown.

In respondents of the 2021 questionnaire in which data regarding actual body weight measurements were obtained from 2019 and 2021, the amount of change in body weight measurements (ΔBW, kg) was stratified into five groups; we examined the reliability of ΔBW against the questionnaire responses (1: “loss of ≥3 kg” to 5: “gain of ≥3 kg”). ΔBW was stratified according to two versions of the criteria as follows: (1) 1: ΔBW < −3, 2: −3 ≤ ΔBW < −1, 3: −1 ≤ ΔBW < +1, 4: +1 ≤ ΔBW < +3, and 5: +3 ≤ ΔBW; and (2) 1: ΔBW < −4.5, 2: −4.5 ≤ ΔBW < 1.5, 3: −1.5 ≤ ΔBW < +1.5, 4: +1.5 ≤ ΔBW < +4.5, and 5: +4.5 ≤ ΔBW. Furthermore, the sensitivity and specificity of the five groups were calculated for the questionnaire responses and ΔBW, and the median ΔBW (25–75 percentile) of each questionnaire response was determined.

### Statistical analysis

Continuous variables other than age were expressed as mean values and standard deviations. The ages were aggregated as mean values and the 25 and 75 percentiles. The reliability of the data regarding the weight gain in the responses to the 2021 questionnaire as a proxy for the result from the actual body weight measurements that were obtained in 2019 and 2021 was evaluated as the agreement using Cronbach’s α coefficient after discretization of the measurement result of the body weight.

In categorical data analyses, the reference group was assigned to a category considered the healthiest or containing the most individuals. The association with weight gain was evaluated by the odds ratio (OR) and 95% confidence interval (CI). The associations were prioritized to be reported using the p values obtained via Fisher’s exact tests. We conducted a binomial logistic regression analysis in which the logistic of the weight gain (responses were dichotomized as explained in the *Study design* section) was regressed onto independent variables with the four highest OR in Fisher’s exact test. We calculated the p value, OR, and 95% CI.

The association of changes in each of the items in sections (3)–(6) with the weight gain was evaluated as the ratio of the McNamar’s chi-square statistics, in which the numerator was the chi-square statistic on the weight gain group and the denominator was the statistic on all participants. Each chi-square statistic was obtained from the contingency table of the dichotomized changes in the factor. The null distribution of the ratio for each factor was obtained as the empirical cumulative distribution function (ECDF) with permutations of the labels (Yes/No) of “weight gain.” For each of the factors, a cutoff value was determined among the thresholds between the adjacent categories as the ECDF returned the extreme value for the observed data.

The distance between the lifestyle factors was defined as the Cramer’s V statistic calculated from the observed data, and clustering analysis among the factors was conducted using the Ward’s hierarchical agglomerative algorithm on the distance.

Statistical analyses were conducted using R version 4.0.3, developed by the R Development Core Team [8].

## Results

### Participant characteristics

Responses were obtained from 3,721 participants (21.3 years old on average) in 2020 and 3,059 participants (21.8 years old on average) in 2021. The WG group that included participants who responded to a weight gain of ≥3 kg during the mild lockdown compared with before the mild lockdown consisted of 224 (6.0%) and 290 (9.5%) students in 2020 and 2021, respectively; compared with 2020, the frequency of students responding to a weight gain of ≥3 kg was significantly higher in 2021 (p < 0.01). In both years, there was a higher number of male students in the WG group than in the non-WG group, and body height, body weight, and body mass index were higher in the WG group (p < 0.05). In both years, compared with the non-WG group, a higher proportion of students in the WG group responded that they experienced a major change in their overall lifestyle (p < 0.01, Table 1).

**Table 1.**
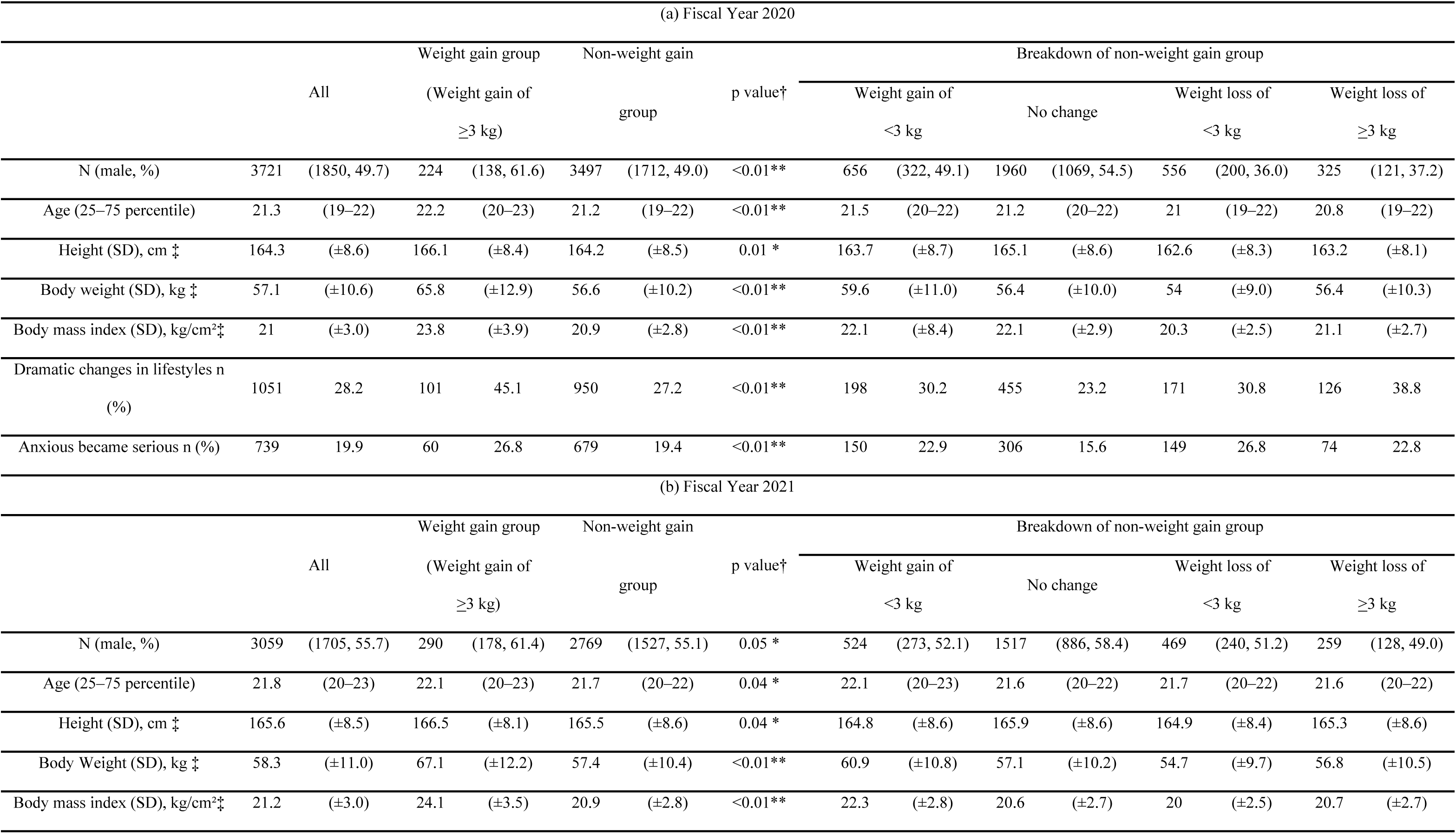
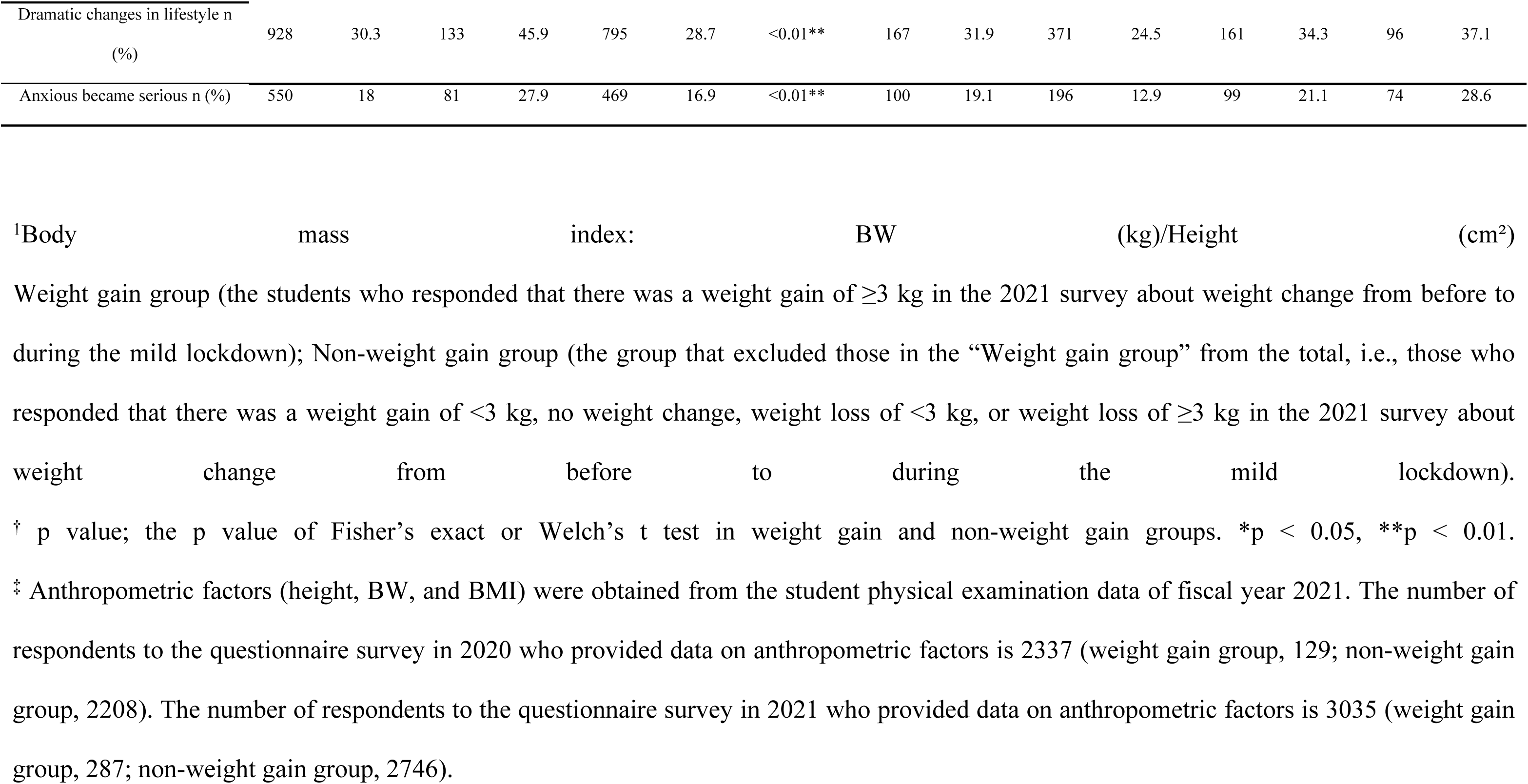
Participant characteristics. (a) Participants from the fiscal year 2020; (b) Participants from the fiscal year 2021.

Regarding the 1,783 students who participated in the 2021 questionnaire and for whom actual body weight measurements were obtained during the 2019 and 2021 medical examinations, we evaluated the amount of change between the questionnaire response and actual body weight measurement (S2 Fig). Regarding the sensitivity and specificity of ΔBW stratified into five groups and questionnaire responses, for weight gain of ≥3 kg and ΔBW of ≥4.5 kg, sensitivity was 0.47 and specificity was 0.95, whereas for ΔBW of ≥3 kg, the values were 0.37 and 0.97, respectively. The median ΔBW was +4.6 (+2.9 to +6.9) kg in the WG group and −0.5 (−2.4 to +1.6) kg in the non-WG group. (The confusion matrix and Cronbach α are shown in S1 Table).

### Clustering analysis of the relationship among changing lifestyle factors during the mild lockdown

To identify the groups of lifestyle factors in which the changes during mild lockdown for all participants (irrespective of body weight change) in 2021 were mutually dependent, we conducted clustering analyses. Because we used Cramer’s V statistic as a measure of the distance between the factors, non-linear and non-rank-ordering associations were included in our findings. As a result, we observed two major groups (groups A and B) of lifestyle factors as well as two and three subgroups of factors under groups A (groups A-a–A-b) and group B (groups B-a–B-c), respectively. The group A included “changes in the time spent at home” (red) and “changes in game time,” “sleeping rhythm,” and “breakfast” (orange). The group B included “changes in smoking” (green), “changes in alcohol consumption” (blue), and other changes (part-time job, club activities, eating out, drinking, and smoking) (purple) (Fig 1).

**Fig 1.**
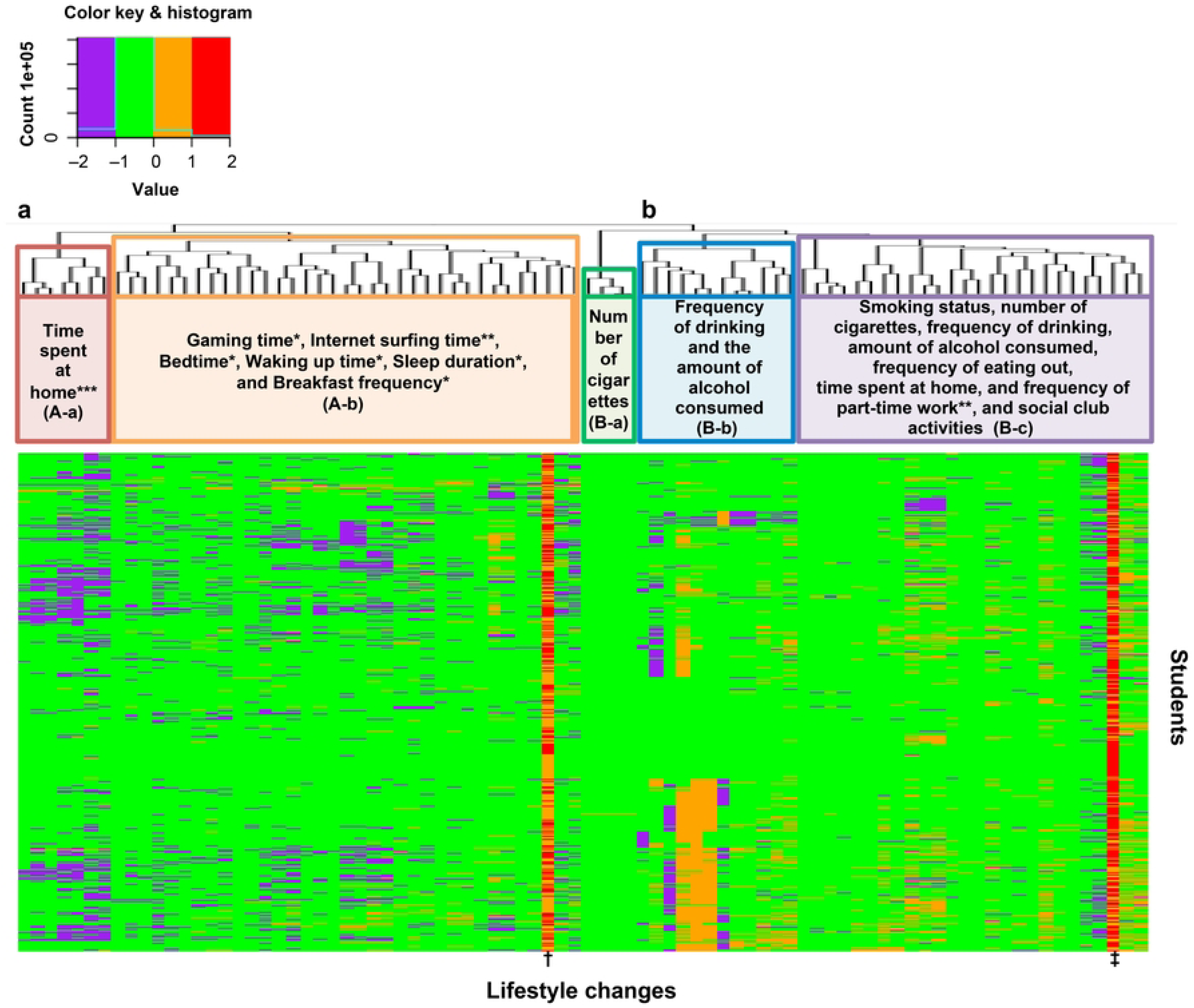
Heatmap showing the clustering of lifestyle changes before and during the mild lockdown. The vertical axis represents students, and the horizontal axis represents each cutoff for lifestyle changes. Purple indicates a change from above to below the cutoff. Light green indicates no change (i.e., above to above or below to below the cutoff). Orange indicates a change from below to above the cutoff. *: Lifestyle factors found to be associated with weight gain using Fisher’s exact test across all students; **: Lifestyle factors found to be associated with weight gain using Fisher’s exact test across male students; ***: Lifestyle factors found to be associated with weight gain using Fisher’s exact test across female students; †: sex, ‡: absence of dramatic lifestyles changes. Regarding sex, orange represents males and red represents females. For overall life change, orange indicates dramatic change and red indicates no dramatic change.

### Relationship between a weight gain of ≥3 kg and each lifestyle factor during the mild lockdown using Fisher’s exact test

The following factors were found to be associated with a ≥3 kg weight gain: OR (95% CI), p value from Fisher’s exact test of the 2021 questionnaire data: breakfast frequency of ≤1 time/week (1.39 [1.01, 1.91], p = 4.7 × 10^−2^), dining out ≥4 times/week (2.16 [1.40, 3.32], p = 8.7 × 10^−4^), bedtime at or after 2 am (1.57 [1.13, 2.20], p = 7.4 × 10^−3^), waking up at or after 10 am (1.43 [1.00, 2.04], p = 4.9 × 10^−2^), sleep duration of ≥9 h (1.74 [1.01, 2.98], p = 4.8 × 10^−2^), and gaming time of ≥4 h/day (2.26 [1.45, 3.47], p = 2.4 × 10^−4^) (Fig 2).

**Fig 2.**
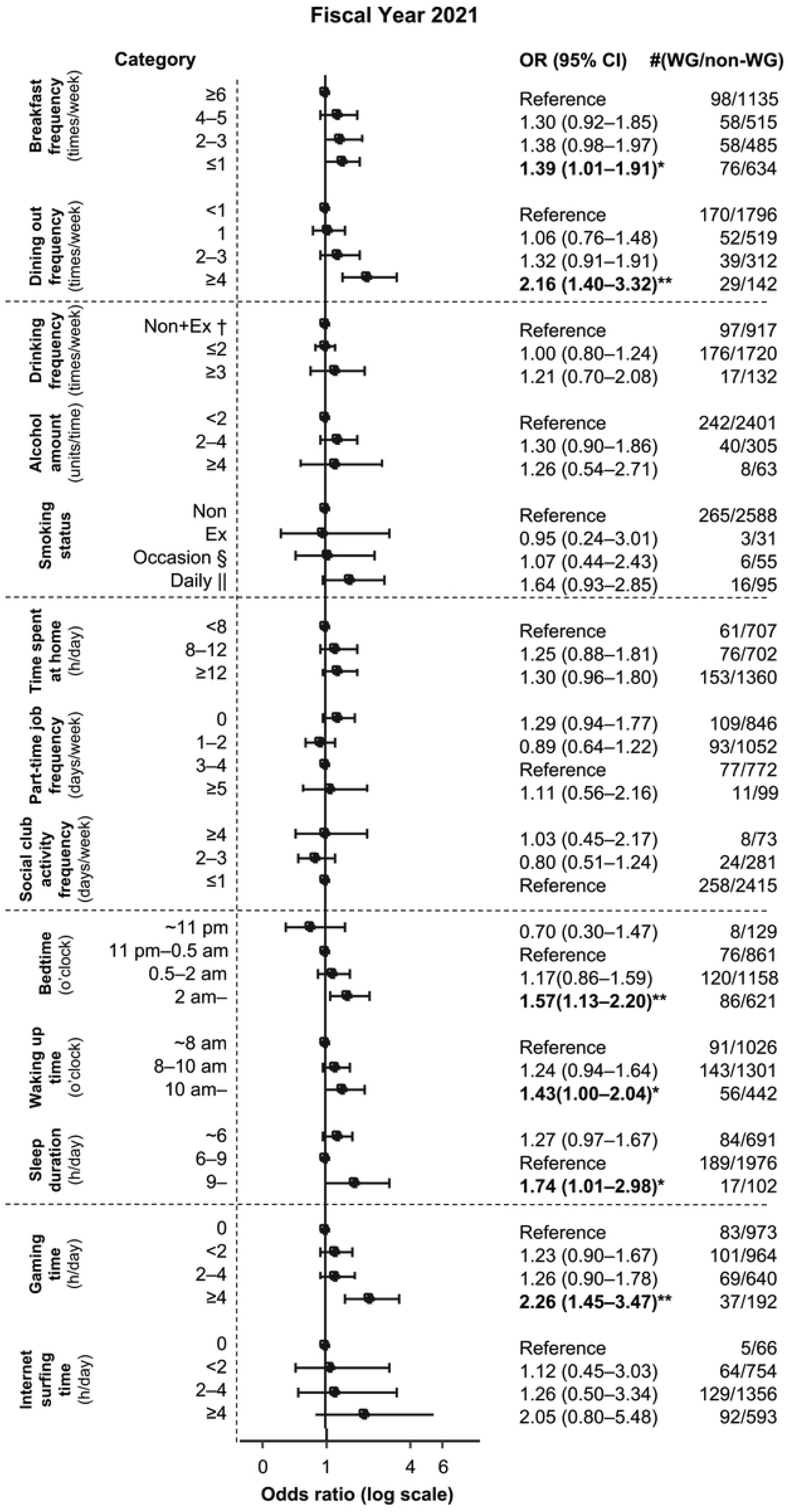
Relationship between weight gain of ≥3 kg and each lifestyle factor during the mild lockdown in all students. *p value < 0.05; **p value < 0.01; †Non: Nondrinker or nonsmoker; ‡Ex: ex-drinker or ex-smoker; §Occasion: occasional smoker; ||Daily: daily smoker. OR: Odds ratio; WG: Weight gain group (the student group who responded that there was a weight gain of ≥3 kg in the 2021 survey about weight change from before to during the mild lockdown); Non-WG: Non-weight gain group (the student group that excluded those in the “Weight gain group” from the total, i.e., those who responded that there was a weight gain of <3 kg, no weight change, weight loss of <3 kg, or weight loss of ≥3 kg in the 2021 survey about weight change from before to during the mild lockdown). Error bars indicate 95% confidence intervals.

Furthermore, in the analysis according to sex, the following factors were found to be associated with a ≥3 kg weight gain in males: dining out ≥4 times/week (2.00 [1.22, 3.27], p = 7.4 × 10^−3^), part-time work frequency of 0 times/week (1.52 [1.02, 2.30], p = 4.8 × 10^−2^), bedtime at or after 2 am (1.64 [1.06, 2.53], p = 2.5 × 10^−2^), waking up between 8 and 10 am (1.47 [1.02, 2.13], p = 3.9 × 10^−2^), waking up at or after 10 am (1.73 [1.09, 2.76], p = 2.5 × 10^−2^), sleep duration of ≥9 h (2.23 [1.10, 4.37], p = 2.8 × 10^−2^), gaming time of ≥4 h/day (2.30, [1.34, 3.93], p = 1.8 × 10^−3^), and internet surfing time of ≥4 h/day (3.00, [1.03, 9.40], p = 3.4 × 10^−2^) (Fig 3A). In females, the factor associated with a ≥3 kg weight gain was time spent at home of ≥12 h/day (1.95 [1.13, 3.39], p = 1.3 × 10^−2^) only (Fig 3B).

**Fig 3.**
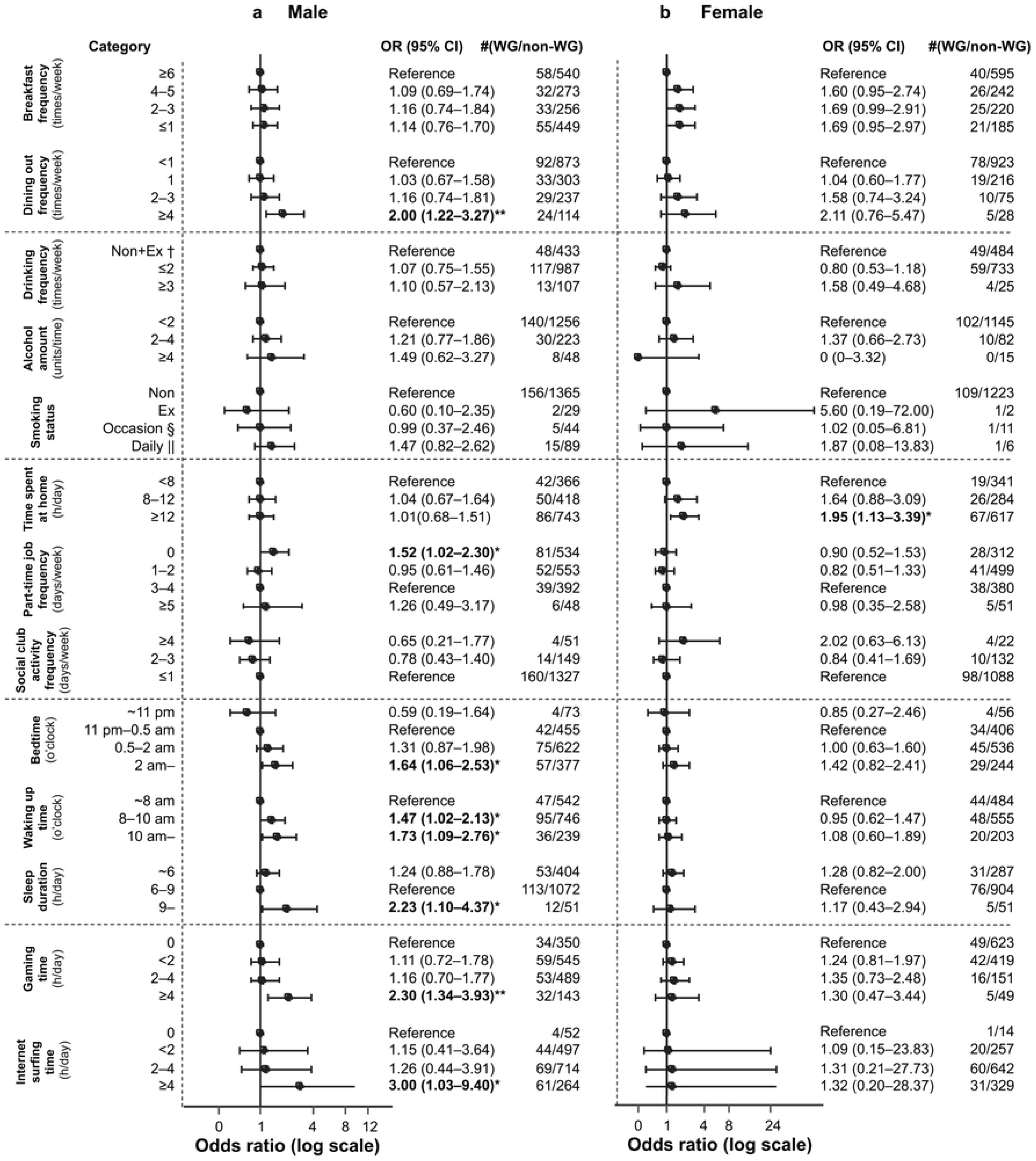
Relationship between weight gain of ≥3 kg and each lifestyle factor during the mild lockdown according to sex. (a) Relationship between weight gain of ≥3 kg and each lifestyle factor during the mild lockdown in male students; (b) the relationship between weight gain of ≥3 kg and each lifestyle factor during the mild lockdown in female students *p value < 0.05; **p value < 0.01; †Non: Nondrinker or nonsmoker; ‡Ex: ex-drinker or ex-smoker; §Occasion: occasional smoker; ||Daily: daily smoker. OR: Odds ratio; WG: Weight gain group (the student group who responded that there was a weight gain of ≥3 kg in the 2021 survey about weight change from before to during the mild lockdown); Non-WG: Non-weight gain group (the student group that excluded those in the “Weight gain group” from the total, i.e., those who responded that there was a weight gain of <3 kg, no weight change, weight loss of <3 kg, or weight loss of ≥3 kg in the 2021 survey about weight change from before to during the mild lockdown). Error bars indicate 95% confidence intervals.

Overall, in 2021, we found that weight gain in students was associated with skipping breakfast, frequently dining out, night owl sleeping rhythm, and gaming time. In males, weight gain was associated with frequently dining out, part-time job frequency, night owl sleeping rhythm, gaming time, and internet surfing time, whereas in females, it was associated with time spent at home.

### Multivariate analysis

We conducted binary logistic regression of the four items with the highest OR in the Fisher’s exact test. The top items in Fisher’s exact test included dining out ≥4 times/week, sleep duration of ≥9 h, gaming time of ≥4 h, and internet surfing time of ≥4 h. Among these, after adjusting for all other factors, a relationship between weight gain and dining out ≥4 times/week and gaming time of ≥4 h was observed (Table 2).

**Table 2.**
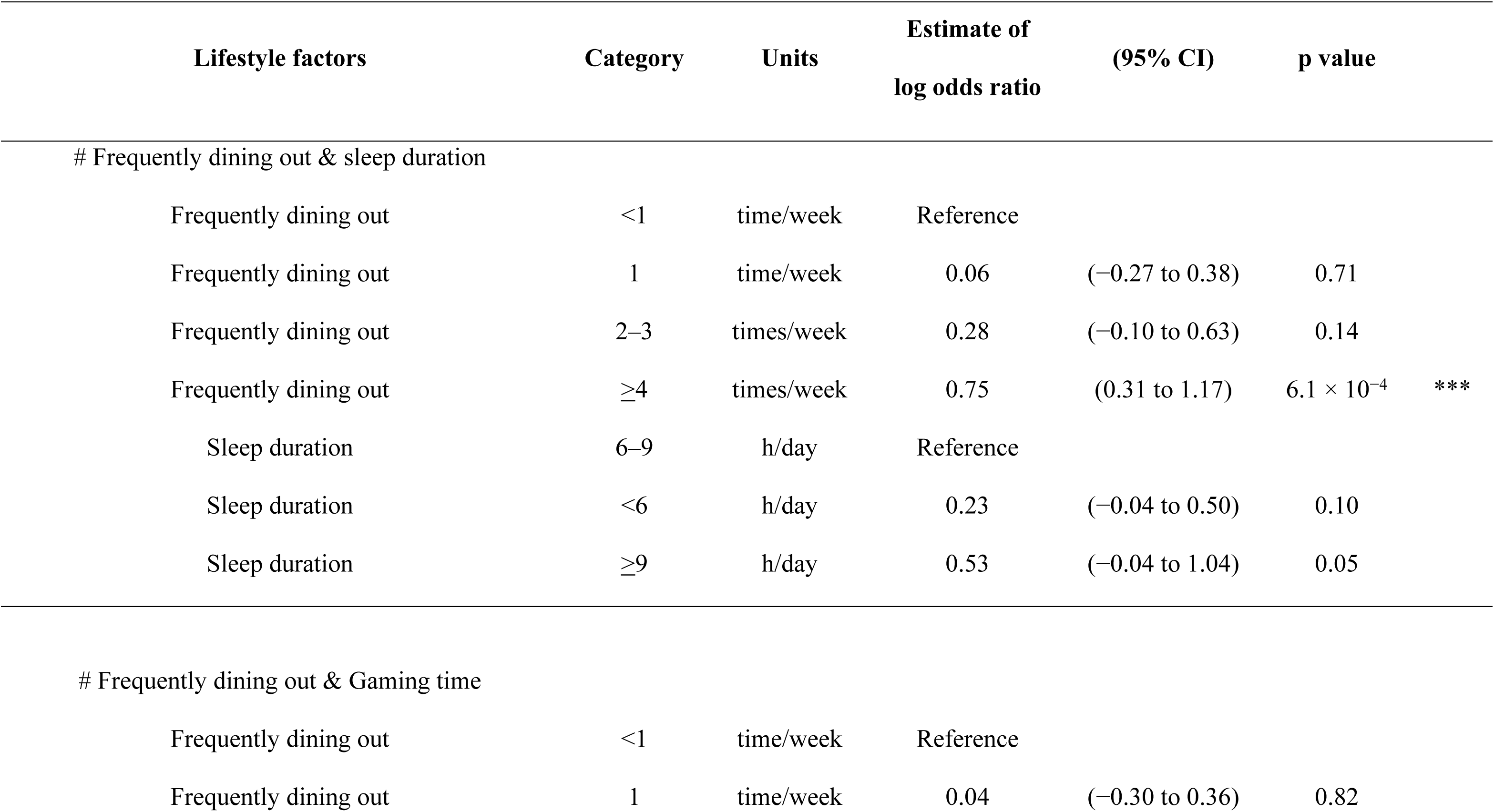
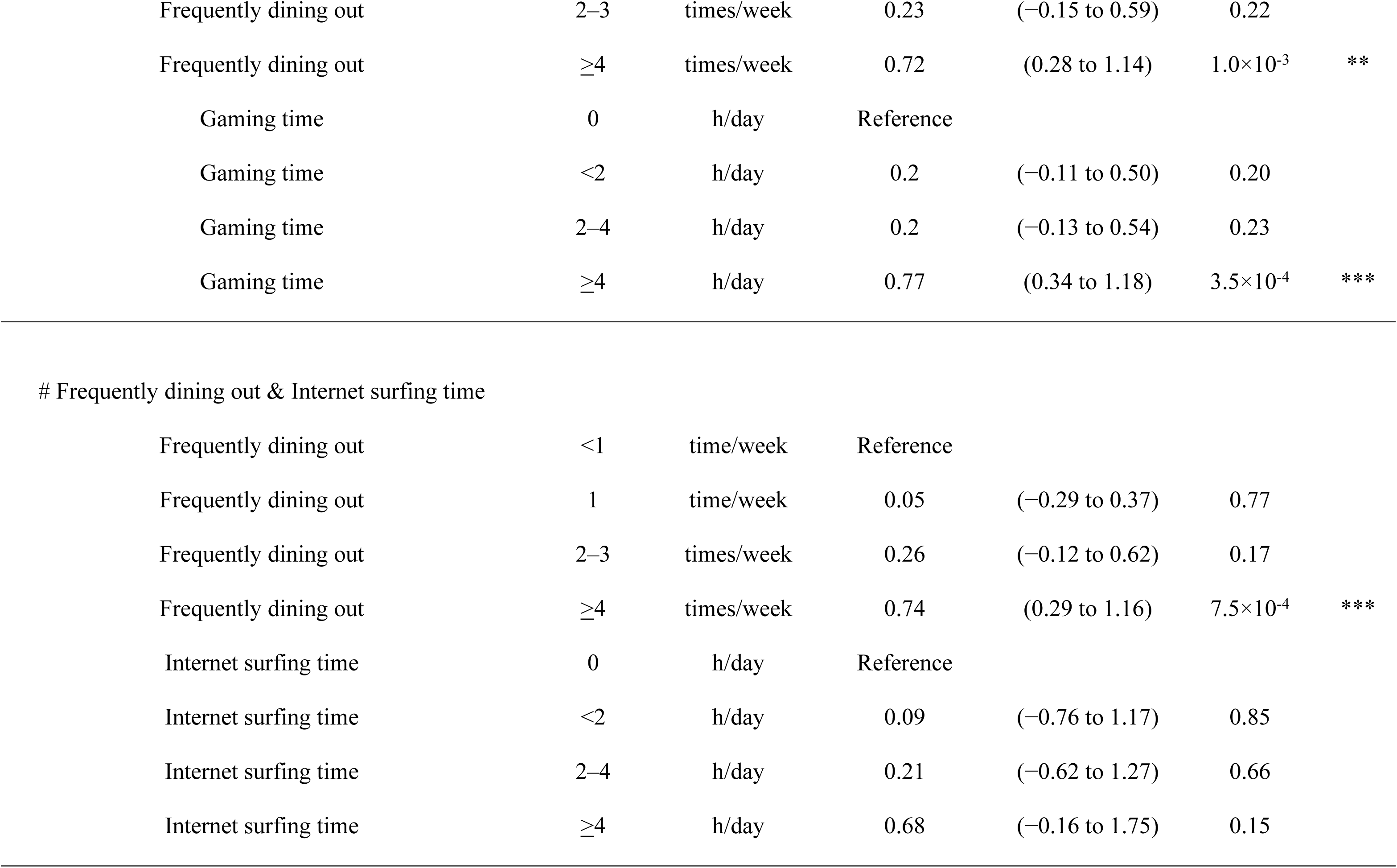
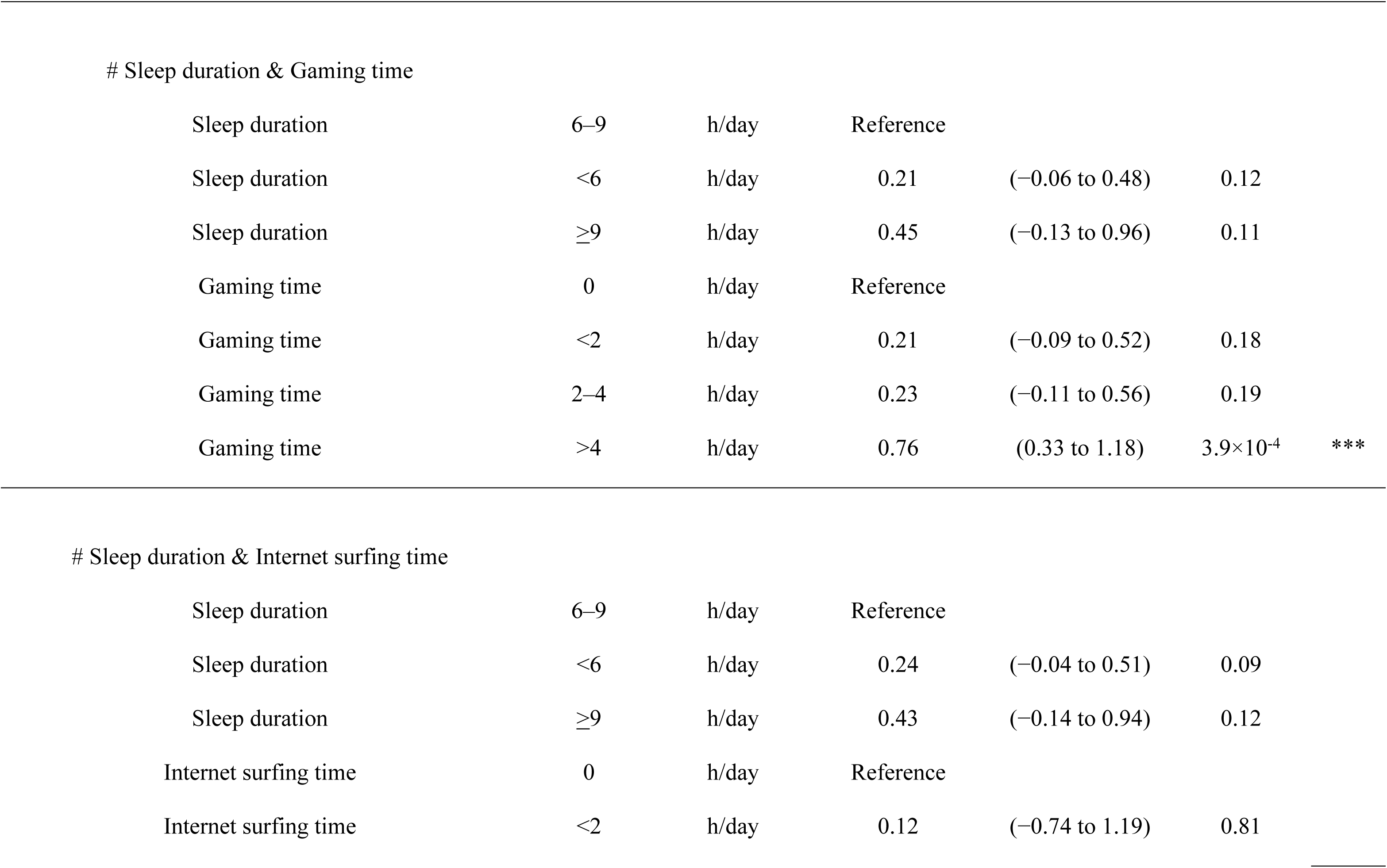
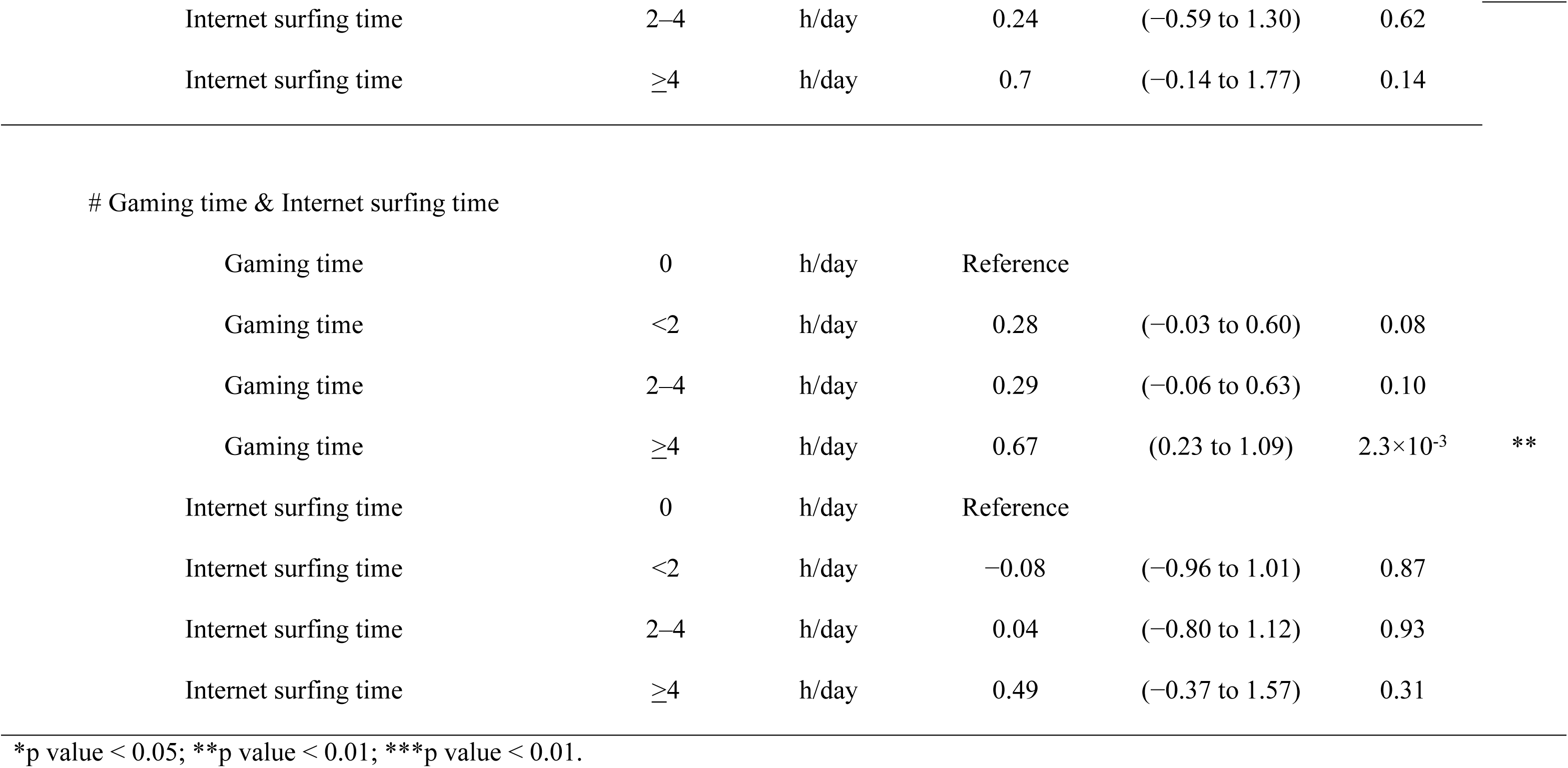
Binary logistic regression analysis for the lifestyle factors associated with a weight gain of >3 kg during mild lockdown in all students.

| Lifestyle factors | Category | Units | Estimate of<br>log odds ratio | (95% CI) | p value |  |
| --- | --- | --- | --- | --- | --- | --- |
| # Frequently dining out & sleep duration |  |  |  |  |  |  |
| Frequently dining out | <1 | time/week | Reference |  |  |  |
| Frequently dining out | 1 | time/week | 0.06 | (−0.27 to 0.38) | 0.71 |  |
| Frequently dining out | 2–3 | times/week | 0.28 | (−0.10 to 0.63) | 0.14 |  |
| Frequently dining out | ≥4 | times/week | 0.75 | (0.31 to 1.17) | $6.1 \times 10^{-4}$ | *** |
| Sleep duration | 6–9 | h/day | Reference |  |  |  |
| Sleep duration | <6 | h/day | 0.23 | (−0.04 to 0.50) | 0.10 |  |
| Sleep duration | ≥9 | h/day | 0.53 | (−0.04 to 1.04) | 0.05 |  |
| # Frequently dining out & Gaming time |  |  |  |  |  |  |
| Frequently dining out | <1 | time/week | Reference |  |  |  |
| Frequently dining out | 1 | time/week | 0.04 | (−0.30 to 0.36) | 0.82 |  |
| Frequently dining out | 2–3 | times/week | 0.23 | (–0.15 to 0.59) | 0.22 |  |
| Frequently dining out | ≥4 | times/week | 0.72 | (0.28 to 1.14) | 1.0×10<sup>–3</sup> | ** |
| Gaming time | 0 | h/day | Reference |  |  |  |
| Gaming time | <2 | h/day | 0.2 | (–0.11 to 0.50) | 0.20 |  |
| Gaming time | 2–4 | h/day | 0.2 | (–0.13 to 0.54) | 0.23 |  |
| Gaming time | ≥4 | h/day | 0.77 | (0.34 to 1.18) | 3.5×10<sup>–4</sup> | *** |
| Frequently dining out & Internet surfing time |  |  |  |  |  |  |
| Frequently dining out | <1 | time/week | Reference |  |  |  |
| Frequently dining out | 1 | time/week | 0.05 | (–0.29 to 0.37) | 0.77 |  |
| Frequently dining out | 2–3 | times/week | 0.26 | (–0.12 to 0.62) | 0.17 |  |
| Frequently dining out | ≥4 | times/week | 0.74 | (0.29 to 1.16) | 7.5×10<sup>–4</sup> | *** |
| Internet surfing time | 0 | h/day | Reference |  |  |  |
| Internet surfing time | <2 | h/day | 0.09 | (–0.76 to 1.17) | 0.85 |  |
| Internet surfing time | 2–4 | h/day | 0.21 | (–0.62 to 1.27) | 0.66 |  |
| Internet surfing time | ≥4 | h/day | 0.68 | (–0.16 to 1.75) | 0.15 |  |
| Sleep duration & Gaming time |  |  |  |  |  |  |
| Sleep duration | 6–9 | h/day | Reference |  |  |  |
| Sleep duration | <6 | h/day | 0.21 | (–0.06 to 0.48) | 0.12 |  |
| Sleep duration | ≥9 | h/day | 0.45 | (–0.13 to 0.96) | 0.11 |  |
| Gaming time | 0 | h/day | Reference |  |  |  |
| Gaming time | <2 | h/day | 0.21 | (–0.09 to 0.52) | 0.18 |  |
| Gaming time | 2–4 | h/day | 0.23 | (–0.11 to 0.56) | 0.19 |  |
| Gaming time | >4 | h/day | 0.76 | (0.33 to 1.18) | 3.9×10<sup>–4</sup> | *** |

Sleep duration & Internet surfing time
|  |  |  |  |  |  |
| --- | --- | --- | --- | --- | --- |
| Sleep duration | 6–9 | h/day | Reference |  |  |
| Sleep duration | <6 | h/day | 0.24 | (–0.04 to 0.51) | 0.09 |
| Sleep duration | ≥9 | h/day | 0.43 | (–0.14 to 0.94) | 0.12 |
| Internet surfing time | 0 | h/day | Reference |  |  |
| Internet surfing time | <2 | h/day | 0.12 | (–0.74 to 1.19) | 0.81 |
| Internet surfing time | 2–4 | h/day | 0.24 | (–0.59 to 1.30) | 0.62 |  |
| Internet surfing time | ≥4 | h/day | 0.7 | (–0.14 to 1.77) | 0.14 |  |
| # Gaming time & Internet surfing time |  |  |  |  |  |  |
| Gaming time | 0 | h/day | Reference |  |  |  |
| Gaming time | <2 | h/day | 0.28 | (–0.03 to 0.60) | 0.08 |  |
| Gaming time | 2–4 | h/day | 0.29 | (–0.06 to 0.63) | 0.10 |  |
| Gaming time | ≥4 | h/day | 0.67 | (0.23 to 1.09) | 2.3×10<sup>–3</sup> | ** |
| Internet surfing time | 0 | h/day | Reference |  |  |  |
| Internet surfing time | <2 | h/day | –0.08 | (–0.96 to 1.01) | 0.87 |  |
| Internet surfing time | 2–4 | h/day | 0.04 | (–0.80 to 1.12) | 0.93 |  |
| Internet surfing time | ≥4 | h/day | 0.49 | (–0.37 to 1.57) | 0.31 |  |

290 \*p value < 0.05; \*\*p value < 0.01; \*\*\*p value < 0.01.

In terms of sex differences, the top four items were identified from Fisher’s exact test for male students; after adjusting for all other factors, a relationship between weight gain and dining out ≥4 times/week and gaming time of ≥4 h was observed. However, the relationship with gaming time disappeared when adjusted with internet surfing time.

Furthermore, a relationship with weight gain remained only when frequently dining out was adjusted with sleep duration of ≥9 h and when internet surfing time of ≥4 h was adjusted with bedtime after 2 am (S2 Table).

The top four items of the Fisher test for females included dining out ≥4 times/week, occasional smoking, ≥12 h spent at home, social club activities ≥4times/week; even after adjusting ≥12 h spent at home with each of the other factors, a relationship with weight gain was observed (p < 0.05, log OR ≥ 0.62, S3 Table).

Furthermore, items with a high log OR (>0.6) when p value was ≥0.05 included internet surfing time of ≥4 h (adjusted with frequently dining out and sleep duration). In males, this included internet surfing time of ≥4 h (adjusted with frequently dining out and gaming time) and sleep duration ≥9 h (adjusted with gaming time and internet surfing time); in females, this included dining out ≥4 times/week and occasional smoking (combined with all other factors) (Table 2, S2 and S3 Tables).

### Relationship between lifestyle changes and weight gain of ≥3 kg before the mild lockdown and at the time of the survey using ECDF

We examined the association of changes in the lifestyle with the weight gain between before the mild lockdown and at the time of the survey, which has not been analyzed in studies to date. We extracted the ECDF value for changes in each cutoff value for each lifestyle factor in 2021 and for weight gain of ≥3 kg from the ratio of the McNemar chi-square statistics (the strength of lifestyle change before and during the mild lockdown) that we obtained from the overall participants to the McNemar chi-square value (the strength of lifestyle change before and during the mild lockdown) for the WG group. Lifestyle changes with an ECDF closer to 1 were considered to have a high association with a weight change of ≥3 kg (such lifestyle changes become clearer after classification according to the presence or absence of weight gain of ≥3 kg and the extraction of only the WG group). Among each lifestyle factor, we extracted the cutoff for the change with the greatest ECDF and the associated ECDF, the ranking of which is presented in Table 3a. The higher the ECDF (close to 1), the higher is the ranking, indicating lifestyle changes with a high relationship to weight gain of ≥3 kg.

**Table 3.**
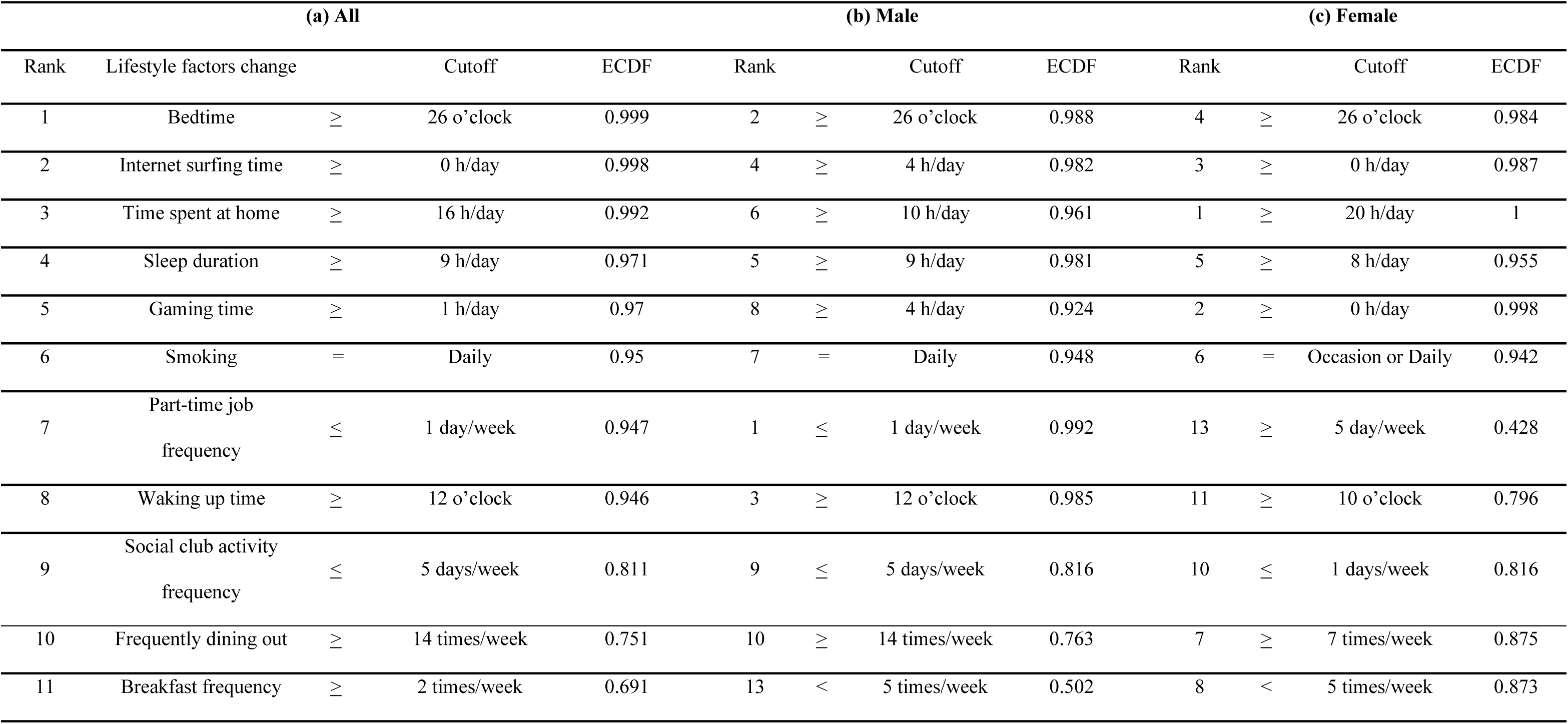
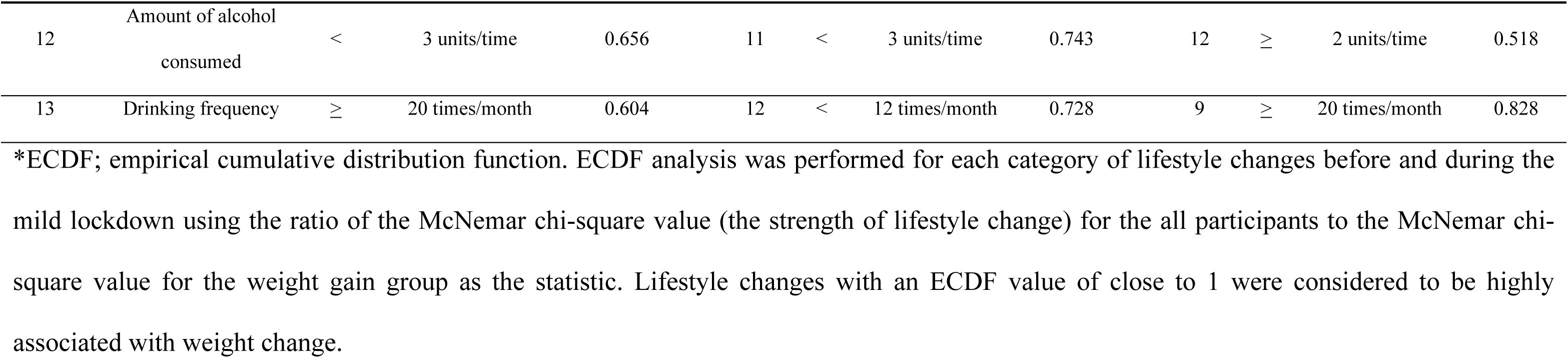
Ranking of ECDF values for the largest value for each lifestyle change factor in 2020 and 2021: (a) the ranking of ECDF values for the largest value for each lifestyle change factor in 2020 and 2021 in all students; (b) the ranking of ECDF values for the largest value for each lifestyle change factor in 2020 and 2021 in male students; (c) the ranking of ECDF values for the largest value for each lifestyle change factor in 2020 and 2021 in female students.

Items with a high ranking ECDF included bedtime at or after 2 am, prolonged internet surfing time > 0 hour, prolonged time spent at home for ≥16 h, prolonged sleeping duration of ≥9 h, and prolonged gaming time of ≥1 h/day (ECDF ≥ 0.96). In terms of sex, although the cutoff value differed, the maximum ECDF for changes in lifestyle factors was high (ECDF ≥ 0.92) (Table 3b, c).

## Discussion

Imposing lockdown during the COVID-19 pandemic is an effective measure for preventing the spread of infection; however, staying at home for a long period can threaten personal health and lead to harmful effects, such as weight gain [4]; in Japan, where a mild lockdown was implemented, there was concern regarding similar issues. Among the students of Nagasaki University in the present survey, the number of students who exhibited weight gain increased with the prolongation of the mild lockdown.

In the present study, a relationship was observed between gaming time and weight gain, and the relationship with changes in gaming, internet surfing, and sleeping times were suggested in overall students and in male students. The population exhibiting a prolonged gaming time accounted for 7.5% of the overall population surveyed (male: 10.3%, female: 4.0%); this population was characteristically male-predominant. Gaming disorder and internet addiction disorder are concepts that have long since been established; however, in recent years, the increase in gaming, particularly among the youth, has drawn attention as an health issue, and in 2019, gaming disorder was proposed as an important new disease concept to be adopted into the ICD-11 of the World Health Organization [9,10]. The prevalence of gaming disorder is reported to be approximately 10%–15% in the youth in Asia, and approximately 1%–10% in the western countries [11] and there is concern that this increase will continue in future. In Japan, it has been reported that with the spread of COVID-19 infection, the incidence of internet gaming disorder has increased, and that young individuals in particular are at high risk of developing this disorder [12].

Reportedly, prolonged time spent on games and the internet causes weight gain owing to factors, such as a higher number of meals consumed during screen time and the increased exposure, preference, and desire to purchase high-calorie foods [13]. In addition, it is inferred that a relative decline in physical activity level is involved. Meanwhile, it has been reported that internet addiction disorder and gaming disorder are deeply associated with mental issues, such as depression, anxiety [14], eating disorder [15], and sleep disturbance [16–21]. In particular, it has been suggested that sleep disturbance disrupts the circadian rhythm and is associated with obesity and hepatic and intestinal diseases caused by the dysregulation of the physiological functions of hepatocytes, gastrointestinal cells, and adipocytes [22,23]. In studies that evaluated the health status of adolescents and youth in the COVID-19 pandemic, game usage and gaming disorder as well as smartphone addiction were reported to exert an adverse effect on health leading to disorders, such as overweight and weight gain via sleep deficit [24–26].

The present study suggested that time spent at home is strongly associated with weight gain, particularly in females. The Ministry of Health, Labour and Welfare of Japan has reported that among adults in their 20 s, 8.3% of men and 13.9% of women responded “a lack of a place or facility” as the reason preventing them from exercising regularly, whereas 5.8% of men and 18.3% of women responded “dislike of exercise” as the reason [1]. Compared with men, women in their 20 s socialize less owing to the increased time spent at home and have less access to places and facilities where they can exercise, which can easily reduce the physical activity level.

Frequently dining out was another factor related with weight gain based on multivariate analysis in this study. However, the ECDF ranking of frequently dining out is low; therefore, it could cause weight gain independently of lifestyle changes voluntarily imposed owing to COVID-19. In our questionnaire survey, although there were students who habitually dined out daily, they were few in number, accounting for only 2.7% and 1.8% before and during the mild lockdown, respectively (data not shown). In a report of Japanese adults in 2015, the proportion of obese individuals accounted for 40.0% of the population who dined out at least twice a day and 22.7%–28.9% of the population who dined out less than twice a day [27], suggesting a relationship between frequently dining out and weight gain from prior to the COVID-19 pandemic.

The present study has several limitations. First, it is a study based on questionnaire responses, and there is no objective data, such as height and weight of all participants; therefore, biases attributed to self-reporting as well as the subject sample being limited to students who provided consent are both possible. Second, the study includes students of a single Japanese institution, and it is possible that generalization will be difficult. To investigate a causal relationship, a prospective study is required in future.

## Conclusions

In the present study, we investigated lifestyle factors that influence weight gain in university students in a particular environment, i.e., the mild lockdown, and confirmed a relationship with factors, such as gaming time, prolonged time spent at home, and night owl sleeping rhythm. Future studies should be conducted to identify the role of early intervention and provide further insights into the conclusions of the study.

## Data Availability

The data that support the findings of this study are available on request from the corresponding author. The data are not publicly available due to privacy reasons.

## Acknowledgments

We thank Mr. Masaki Miwa, Ms. Hiroko Yamamoto, and Ms. Chikako Yamaura at the Diabetes care support center, Nagasaki University for their assistance in conducting the survey, and Ulatus (www.ulatus.jp/) for language editing.

## Institutional review board statement

The study was conducted in accordance with the Declaration of Helsinki, and approved by the ethical review board of Nagasaki University (approval number: 20062604).

## Informed consent statement

Informed consent was obtained from all subjects involved in the study. Written informed consent has been obtained from the patients to publish this paper.

## Data availability statement

The data presented in this study are available on request from the corresponding author. The data are not publicly available due to prevent misuse and interpretation of data.

## Author Contributions

Conceptualization, N.A., K.A., H.A., and M.K.; methodology, formal analysis, S.M.; investigation, K.A., resources, T.N. and M.K.; writing - original draft preparation, S.M., N.A., K.A. and M.K.; funding acquisition, A.K. (Atsushi Kawakami) and M.K.; writing - review and editing, N.A.; visualization, S.M.; supervision, N.A., A.K. and M.K; project administra-tion, M.K. All authors have read and agreed to the published version of the manuscript.

## Supporting information

**S1 Fig. Study flow chart.** In 2020, of the 6,065 students who received medical examinations, 2,344 did not provide consent for participation or a complete response. In 2021, of the 6,675 students who received medical examinations, 3,616 did not provide consent for participation. We analyzed the data from the questionnaire with 3,721 and 3,059 students in 2020 and 2021, respectively. After 1,783 students of participants in 2021 received the 2019 medical examinations, we evaluated the amount of change between the questionnaire response and actual body weight measurement. In the 2020 medical examination, weight measurements were not conducted for infection control.

**S2 Fig. Self-reported weight change in the fiscal year 2021 questionnaire and the amount of actual weight change based on student health examination data from fiscal year 2019 to 2021.** ΔBW: actual weight change based on student health examination data from fiscal year 2019 to 2021 (kg); Weight gain group: the student group who responded that there was a weight gain of ≥3 kg in the 2021 survey about weight change from before to during the mild lockdown; Non-weight gain group: the student group that excluded those in the “Weight gain group” from the total, i.e., those who responded that there was a weight gain of <3 kg, no weight change, weight loss of <3 kg, or weight loss of ≥3 kg in the 2021 survey about weight change from before to during the mild lockdown. Error bars range from (first quartile − quartile range) × 1.5 to (third quartile + quartile range) × 1.5.

**S1 Table. Confusion matrix and Cronbach’s α of self-reported weight change in the fiscal year 2021 questionnaire and the amount of actual weight change based on student health examination data from fiscal year 2019 to 2021.**

**S2 Table. Binary logistic regression analysis for the lifestyle factors associated with a weight gain of >3kg during the mild lockdown in male students.**

**S3 Table. Binary logistic regression analysis for the lifestyle factors associated with a weight gain of ≥3kg during the mild lockdown in female students.**

## Notes

### Competing Interest Statement

The authors have declared no competing interest.

